# Feature-Engineering Strategies for EEG-Based ADHD Classification in Children: A Controlled Benchmark of Expert, LLM-Guided and Full-Space Comparators

**DOI:** 10.64898/2026.10.06.26364825

**Authors:** Faezeh Rohani, Hasan Alhatab, Mohamed Jarrud, Nour Salous, Salman Kasimoğlu, Andreas Mueller

## Abstract

EEG-based machine-learning studies of attention-deficit/hyperactivity disorder (ADHD) are vulnerable to optimistic performance estimates when feature selection or model choice is informed by data outside the training fold. We compared expert-reconstructed, large language model (LLM)-guided and full-space feature strategies under leakage-controlled subject-level validation in a frozen cohort of 77 children (45 ADHD, 32 controls) represented by 1,185 gamma-free EEG features. Four classifiers (SVM, GMM, random forest [RF] and XGBoost) were evaluated with nested leave-one-subject-out cross-validation (LOOCV), with all data-dependent operations restricted to outer-training subjects. Across 100 repetitions of stratified 10-fold cross-validation, RF/expert achieved mean accuracy 62.8% (SD 3.5%; empirical 95% interval 55.8%–69.5%), mean balanced accuracy 62.3% and mean AUC-ROC 0.618. The corresponding LOOCV point estimate was 71.4% accuracy and 71.0% balanced accuracy (AUC-ROC 0.673). GMM/expert achieved the highest AUC-ROC (0.699), indicating metric-dependent model ranking. The expert-reconstructed strategy achieved the highest accuracy and balanced accuracy for every classifier, whereas the LLM-guided strategy showed no consistent advantage over the full feature space. Pairwise classifier differences were not significant after Holm correction. A pre-specified 200 µV artifact gate reduced RF/expert accuracy to approximately 55.8%, below the 58.4% no-information rate; this drop may reflect removal of artifact-related signal and/or reduced epoch counts leading to noisier feature estimates. These findings support cautious benchmarking and external validation rather than claims of algorithmic or LLM superiority.

## 1. Introduction

Attention-deficit/hyperactivity disorder (ADHD) is a common neurodevelopmental condition characterized by persistent inattention and/or hyperactivity-impulsivity that can affect academic, social and family functioning [1]–[5]. Diagnosis remains clinical and relies on developmental history, structured or semi-structured assessment, rating scales and clinician judgment. Objective physiological measures may eventually complement, but not replace, this clinical process.

Electroencephalography (EEG) is attractive for research on ADHD because it is non-invasive, relatively inexpensive, temporally precise and directly measures electrical brain activity. Quantitative EEG studies have repeatedly examined spectral power, theta/beta ratio (TBR), task-related oscillatory changes, coherence and event-related measures [6]–[14]. However, the literature also shows substantial heterogeneity across ADHD phenotypes and across EEG-defined subgroups [4], [13]–[15]. Recent pediatric work has also identified distinct EEG-based time-on-task vigilance trajectories in children with and without ADHD [16]. No single EEG marker has established sufficient sensitivity and specificity to serve as a stand-alone diagnostic test [7]–[11].

Machine learning offers a way to combine many weak signals, but small neurophysiological datasets create a second problem: validation procedures can dominate the apparent performance. Subject identity must remain separated across training and testing and any operation that uses labels or data-dependent thresholds must be fitted without access to the held-out subject [17]–[19]. Feature selection on the complete dataset before cross-validation or selecting the best result from many post-hoc configurations, can substantially overstate generalization. Small samples also produce wide uncertainty around cross-validation estimates [17], [18].

A related challenge is feature-space design. High-dimensional EEG representations can contain hundreds or thousands of correlated variables, yet aggressive data-driven selection on a small cohort risks overfitting [20], [21]. Large language models (LLMs) provide a different form of assistance: they can organize feature metadata according to concepts represented in the scientific literature without being given participant labels. This use is potentially valuable for transparent hypothesis generation, but it does not guarantee that literature-prominent feature families are the most discriminative in a specific dataset.

To minimize these risks, the present study uses a frozen, fold-contained framework in which all data-dependent feature selection and model tuning are restricted to the training data, enabling a controlled comparison of the three feature strategies.

We asked three questions. First, under a common leakage-controlled validation framework, how do the full feature space, a reconstructed expert EEG strategy and LLM-guided feature subsets compare? Second, are the observed differences among SVM, GMM, RF and XGBoost large enough to support claims of classifier superiority? Third, how robust are the leading results to repeated cross-validation, alternative preprocessing choices, calibration analysis and prevalence-aware clinical interpretation?

## 2. Methods

### 2.1 Study Design, Dataset and Participants

This was a secondary analysis of the pediatric EEG/ERP corpus originally collected by Rohani et al. [22] at the Motor Behavior Laboratory, Ferdowsi University of Mashhad, Iran, between July 2019 and February 2020. The source cohort contained 103 children: 49 with ADHD and 54 healthy controls. The source article reports an age range of 7–10 years, whereas accompanying dataset metadata describes 6–10 years; participant-level ages for the frozen analytical cohort were not retained in the project records available for the present analysis, so this discrepancy is reported rather than resolved. The source cohort included 22 girls and 27 boys in the ADHD group and 24 girls and 30 boys in the control group [22].

ADHD diagnoses were based on DSM-5 criteria and clinical screening. Participants with IQ below 75, epilepsy or other neurological disorders or relevant psychiatric comorbidity were excluded in the source study. The source documentation states that all children were medication-free at recording. The source study was approved by the Research Ethics Committee of Islamic Azad University, Lahijan Branch, Iran (IR.IAU.LIAU.REC.1398.013) and the parents of all participants provided written informed consent [22].

The present analysis uses a frozen 77-subject feature matrix containing 45 ADHD and 32 control participants. Relative to the 103-subject source cohort, 4 of 49 ADHD participants (8.2%) and 22 of 54 controls (40.7%) are not represented. Individual-level exclusion reasons were not preserved with sufficient detail to support reason-specific exclusion counts. A repository audit identified 83 cleaned subjects (51 ADHD, 32 controls), including six ADHD recordings that were not part of the frozen 77-subject analytical matrix. These additional recordings were not incorporated because the analytical cohort had already been fixed before the final validation analyses. This discrepancy is treated as a cohort-provenance limitation in the Discussion and a corresponding sensitivity analysis is proposed in Section 3.7.

**Table 1.** Available characteristics of the source and frozen analytical cohorts. Abbreviations: EO, eyes open; EC, eyes closed; VCPT, visual continuous performance task.

| Characteristic | Source cohort | Frozen analytical cohort |
| --- | --- | --- |
| Total N | 103 | 77 |
| ADHD / control | 49 / 54 | 45 / 32 |
| Sex | ADHD: 22F/27M; Control: 24F/30M | 45M / 32F |
| Age | 7–10 y in source article; 6–10 y in dataset metadata | 6–10 y |
| Diagnostic framework | DSM-5 clinical diagnosis | Inherited source diagnosis |
| IQ criterion | Exclude IQ <75 (Raven matrices) | Inherited source criterion |
| Medication | Medication-free at recording | Inherited source status |
| Recording conditions | EO, EC, VCPT/Go-NoGo | EO, EC, VCPT available in frozen matrix |
| Analytical prevalence | 47.6% ADHD | 58.4% ADHD |

The differential attrition is a potential source of selection bias and prevents assuming that the 77-subject cohort is demographically identical to the source cohort.

### 2.2 EEG Recording and Pre-processing

In the source study, EEG was recorded at 500 Hz from 19 electrodes of the international 10–20 system (Fp1, Fp2, F3, F4, C3, C4, P3, P4, O1, O2, F7, F8, T3, T4, T5, T6, Fz, Cz, Pz) with linked-earlobe reference and electrode impedance below 5 kΩ. Resting EEG comprised 5 minutes eyes closed and 5 minutes eyes open. The Go/NoGo task contained 400 trials over approximately 20 minutes [22], [23].

The current analysis used cleaned EDF files produced by the project preprocessing pipeline rather than reprocessing the original raw recordings. In this exported analysis set, non-EEG channels were removed, a 0.5–30 Hz FIR band-pass and 50 Hz notch filter were applied and Infomax independent component analysis was used with an Fp1-based rule for identifying ocular components. Residual amplitude anomalies were annotated in the cleaned EDFs. MNE-Python was used for the scripted EEG workflow [24]. Because the original raw files and complete per-participant ICA logs were not present in the retained repository, ICA removal counts could not be compared between diagnostic groups.

The downstream primary feature extractor did not reject epochs merely because they overlapped inherited artifact annotations. To assess whether this choice influenced performance, a pre-specified sensitivity analysis imposed a 200 µV peak-to-peak epoch gate; this analysis is reported separately in Sections 3.7 and 4.6 and was not adopted into the primary pipeline.

### 2.3 Feature Extraction

Cleaned recordings were segmented into 4-second epochs with 2-second overlap. Features were computed per epoch and then aggregated to one subject-level vector for each recording condition using the mean and standard deviation. The primary matrix contains 197 epoch-level features × 2 aggregations × 3 conditions plus 3 VCPT-specific contrasts, yielding 1,185 features for each of 77 subjects.

Spectral power was estimated using Welch periodograms (nperseg = 512). The implemented bands were delta 0.5–4 Hz, theta 4–8 Hz, alpha 8–13 Hz and beta 13–30 Hz. Gamma features were excluded because the exported analysis files had already been low-pass filtered at 30 Hz; residual energy above that cutoff was not treated as physiological gamma. Approximate entropy and sample entropy used the complete 4-second epoch (m = 2; tolerance = 0.2 × signal SD). Magnitude-squared coherence used nperseg = 128 in the primary analysis. No epoch-level sample was treated as an independent observation in model validation. Detailed per-family channel and condition coverage is provided in Supplementary Table S8.

**Table 2.** Final gamma-free feature families in the 1,185-feature primary matrix.

| Feature family | Number of subject-level features |
| --- | --- |
| Band power and spectral ratios | 774 |
| Entropy / complexity | 162 |
| Theta-beta ratio | 78 |
| Inter-hemispheric coherence | 72 |
| Time-domain statistics | 42 |
| Frontal asymmetry | 36 |
| Hjorth parameters | 18 |
| VCPT-specific contrasts | 3 |
| Total | 1,185 |

### 2.4 Feature-Engineering Strategies

#### 2.4.1 Full Feature Space

The full strategy exposed all 1,185 valid columns to the fold-contained variance, correlation and ANOVA selection pipeline. It served as the high-dimensional, non-LLM baseline.

#### 2.4.2 Reconstructed Expert EEG Strategy

The expert comparator contained 450 available features reconstructed from the EEG categories emphasized in Rohani et al. [22]: theta/beta ratio, EO/EC coherence and available theta/alpha/beta channel-bandpower measures. It is deliberately labeled “expert-reconstructed” rather than “Rohani 113” because the exact 113-variable feature vector used in the original study could not be reproduced from the features available for the present analysis. Arousal-index variables, omission and commission errors, reaction time and true ERP amplitude and latency measures were not available in the primary feature table and were therefore excluded from the reconstructed comparator.

#### 2.4.3 LLM-Guided Strategy

The LLM was used only as a literature-informed organizer of feature metadata. It was not given participant identifiers, diagnostic labels, raw EEG, train/test splits or model performance. Seven candidate masks were registered: VCPT-only (397 features), frontal-all (324), VCPT + frontal + TBR (424), global + asymmetry + coherence (414), entropy + Hjorth + time-domain (222), EO + EC only (788) and VCPT + entropy + TBR (505). For the single “LLM-guided best” strategy, the candidate mask itself was chosen inside the outer-training data by inner cross-validation; it was not selected after viewing all 77 outer predictions.

The project records preserve the information-access restrictions and the resulting feature masks, but not a verifiable LLM provider or model build, interaction timestamp or verbatim original prompt. A reconstructed prompt specification, based on the preserved feature-group definitions and documented information-access constraints, is provided in Supplementary Methods S2.

### 2.5 Leakage-Controlled Subject-Level Validation

Primary performance was estimated with 77-fold leave-one-subject-out cross-validation (LOOCV). For each outer fold, the held-out child was completely excluded from all operations involving labels or data-dependent statistics. Within the remaining training subjects, the pipeline applied zero-variance removal, a training-only correlation filter (absolute Pearson r > 0.95; the lower ANOVA F-score feature was removed), ANOVA SelectKBest, scaling where required and model hyperparameter selection. SVM used k = 25; GMM, RF and XGBoost used k = 20.

These k values were fixed before the final validation runs and held constant across all outer folds; they were not re-optimized against outer-fold outcomes. Inner tuning of k over the range 10–40 was evaluated as a pre-specified sensitivity analysis and was not adopted (Section 3.7).

Inner model selection used stratified 5-fold cross-validation with F1 as the frozen selection metric. For the LLM-guided best strategy, the choice among the seven candidate masks was part of this inner loop. The selected complete pipeline was then refitted on the outer-training set and applied once to the untouched outer-test subject.

Random seeds were deterministic (base seed 42 with fold-specific offsets) and model training used n_jobs = 1 to improve reproducibility. The same outer-fit function was reused for repeated stratified cross-validation. No post-hoc probability threshold optimization was used.

**Figure 1.**
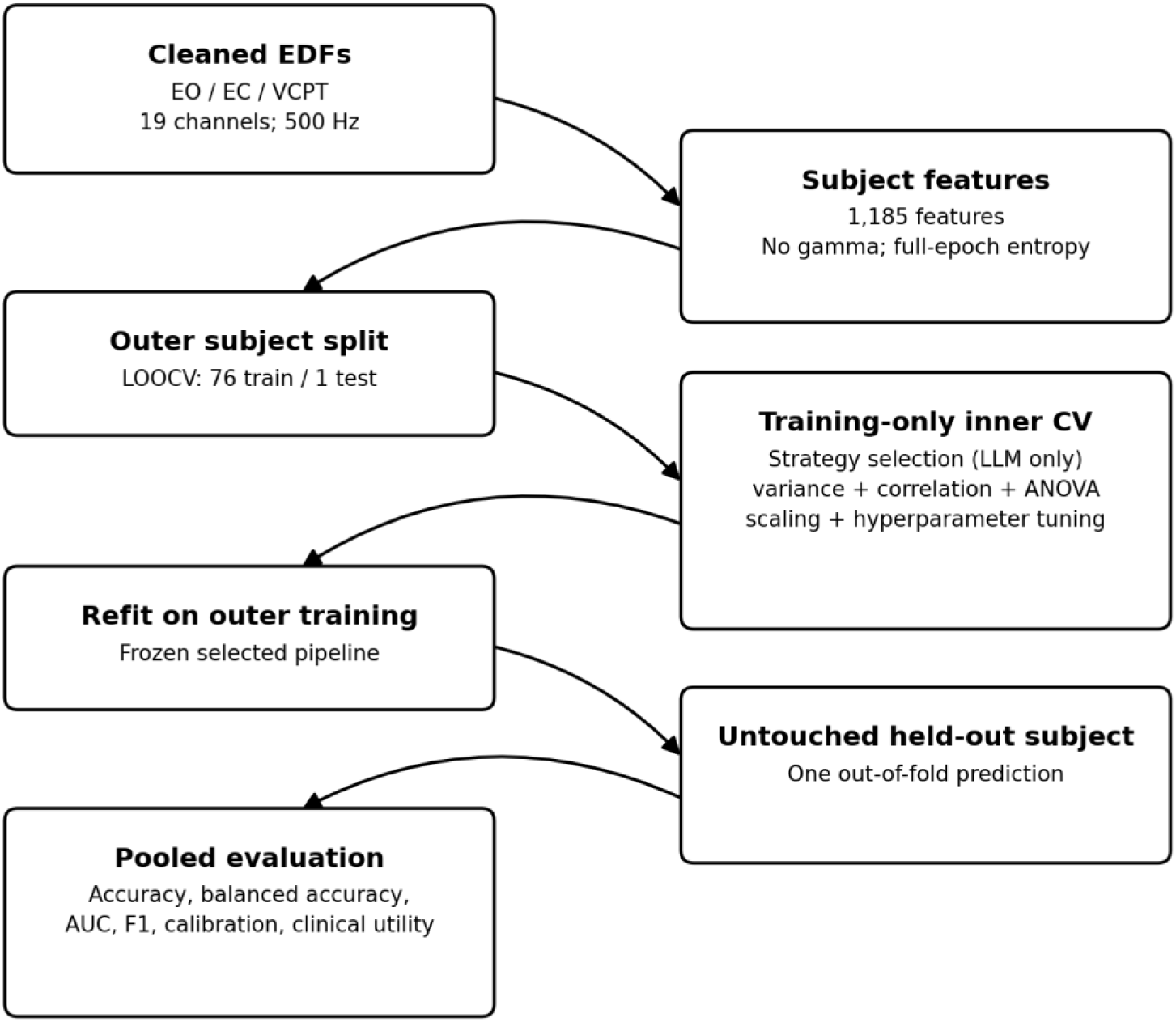
Leakage-controlled subject-level validation. Every label-dependent or data-dependent operation is fitted only within outer-training subjects before prediction of the untouched held-out child.

### 2.6 Classification Models

#### 2.6.1 Support Vector Machine

The SVM analysis used an RBF SVC [25] with ANOVA k = 25, C in {1, 10, 100} and gamma in {scale, 0.01}. The primary tuning path used class_weight = “balanced” with train-only sigmoid calibration. Because this calibration yielded poor specificity and unstable pooled ranking, six alternative scoring constructions were examined using the same outer splits and inner-selected hyperparameters.

The scoring route adopted for reporting is referred to throughout as the uncalibrated standardized-margin route. It refitted the selected RBF SVM with class_weight = None, used the sign of the decision function for class prediction and standardized each held-out margin using the mean and standard deviation of decision values from the corresponding outer-training data before pooled AUC calculation. Because this route was selected after examining six variants and altered the final class-weight setting relative to the frozen inner search, all SVM results are treated as secondary sensitivity evidence and are not used to claim algorithmic superiority.

#### 2.6.2 Gaussian Mixture Model

The GMM was implemented as a class-conditional generative classifier. After the common preprocessing and StandardScaler, eight principal components were retained. Separate diagonal-covariance Gaussian mixtures were fitted for ADHD and control training subjects, with one or two components selected in the inner grid. Classification used normalized class posterior scores including class priors [26].

#### 2.6.3 Random Forest

Random Forest used 250 trees and class_weight = “balanced” [27]. The inner grid varied maximum depth (unrestricted or 5) and minimum samples per leaf (1 or 2), with max_features = “sqrt”. The final model-strategy combination was selected using the pre-specified rule of maximum LOOCV balanced accuracy, followed by AUC-ROC and then fewer candidate features.

#### 2.6.4 XGBoost

XGBoost used binary logistic loss, histogram tree construction, 150 estimators, subsample = 0.8, colsample_bytree = 0.8 and an inner grid over maximum depth {2, 3} and learning rate {0.03, 0.10}. A sensitivity analysis adding scale_pos_weight = 32/45 reduced performance and was not adopted [28].

#### 2.6.5 Linear Reference Model

An elastic-net logistic regression was also run as a secondary linear reference floor. It was not included in the four-model primary comparison. On the expert-reconstructed strategy its accuracy was 57.1% and balanced accuracy 57.0%, below all four nonlinear expert-strategy cells.

### 2.7 Performance Metrics and Statistical Analysis

Balanced accuracy was used as the principal model-selection metric, with pooled out-of-fold AUC-ROC as the first tie-breaker. We also report accuracy, sensitivity, specificity, precision, F1, AUC-PR and confusion-matrix counts. The no-information rate (NIR) was 45/77 = 58.44%, corresponding to always predicting ADHD. Wilson 95% confidence intervals are used for accuracy, sensitivity and specificity where reported. These intervals are descriptive binomial intervals and do not account for dependence among LOOCV predictions arising from overlapping training sets; repeated cross-validation was therefore used as the primary robustness analysis for sampling variability.

Robustness was assessed with repeated stratified 10-fold cross-validation (100 complete repetitions). Each repetition produced exactly one out-of-fold prediction per participant and metrics were calculated on the concatenated predictions for that repetition rather than treating the 1,000 individual folds as independent observations. The distribution across the 100 repeat-level metrics is summarized by the mean, standard deviation, median, quartiles and empirical 2.5th–97.5th percentiles.

Pairwise classifier comparisons used exact McNemar tests on paired out-of-fold class predictions with Holm correction. Label-permutation inference used 1,000 permutations for selected final configurations. The available RF/expert and GMM/expert permutation results are conditional fixed-configuration tests: the selected configuration is frozen and labels are permuted, so the p-values do not correct for prior feature-strategy and hyperparameter selection. This limitation is stated wherever the permutation results are interpreted.

For the primary RF/expert model, calibration was assessed from the 77 out-of-fold probabilities with the Brier score and a logistic calibration model (intercept and slope with model-based 95% confidence intervals). An exploratory decision-curve analysis was performed on the study sample. Because the cohort is enriched for ADHD, decision-curve and predictive-value results are not treated as deployment estimates. PPV and NPV were calculated from the observed sensitivity and specificity at illustrative prevalences of 5%, 10% and 30% and at the study prevalence. Positive and negative likelihood ratios (LR+ and LR−) were also reported.

Reporting was prepared with reference to the TRIPOD+AI statement for prediction-model studies [29].

### 2.8 Sensitivity, Stability and Error Analyses

A sensitivity-analysis suite, with adoption criteria fixed before final model reporting, was used to test whether plausible implementation changes materially altered the conclusions. Evaluated changes included log10 transformation of absolute-power variables, XGBoost class weighting, inner tuning of ANOVA k, changing the inner selection metric from F1 to balanced accuracy, a 200 µV peak-to-peak artifact gate and increasing coherence nperseg from 128 to 500. Changes were adopted only if they met their pre-stated criteria; none was adopted globally. Feature-selection frequencies and error overlap across model-strategy cells were calculated after the primary analysis.

## 3. Results

### 3.1 Cohort and Final Feature Space

The frozen analytical cohort comprised 77 children (45 ADHD, 32 controls), giving an ADHD prevalence of 58.44% and an NIR of 58.44%. The analytical feature table contained 1,185 candidate EEG features with no gamma columns. The three primary strategy sizes were 1,185 features (full), 450 features (expert-reconstructed) and one of seven registered LLM masks selected inside each outer-training fold.

### 3.2 Controlled Feature-Strategy Comparison

**Table 3a.**
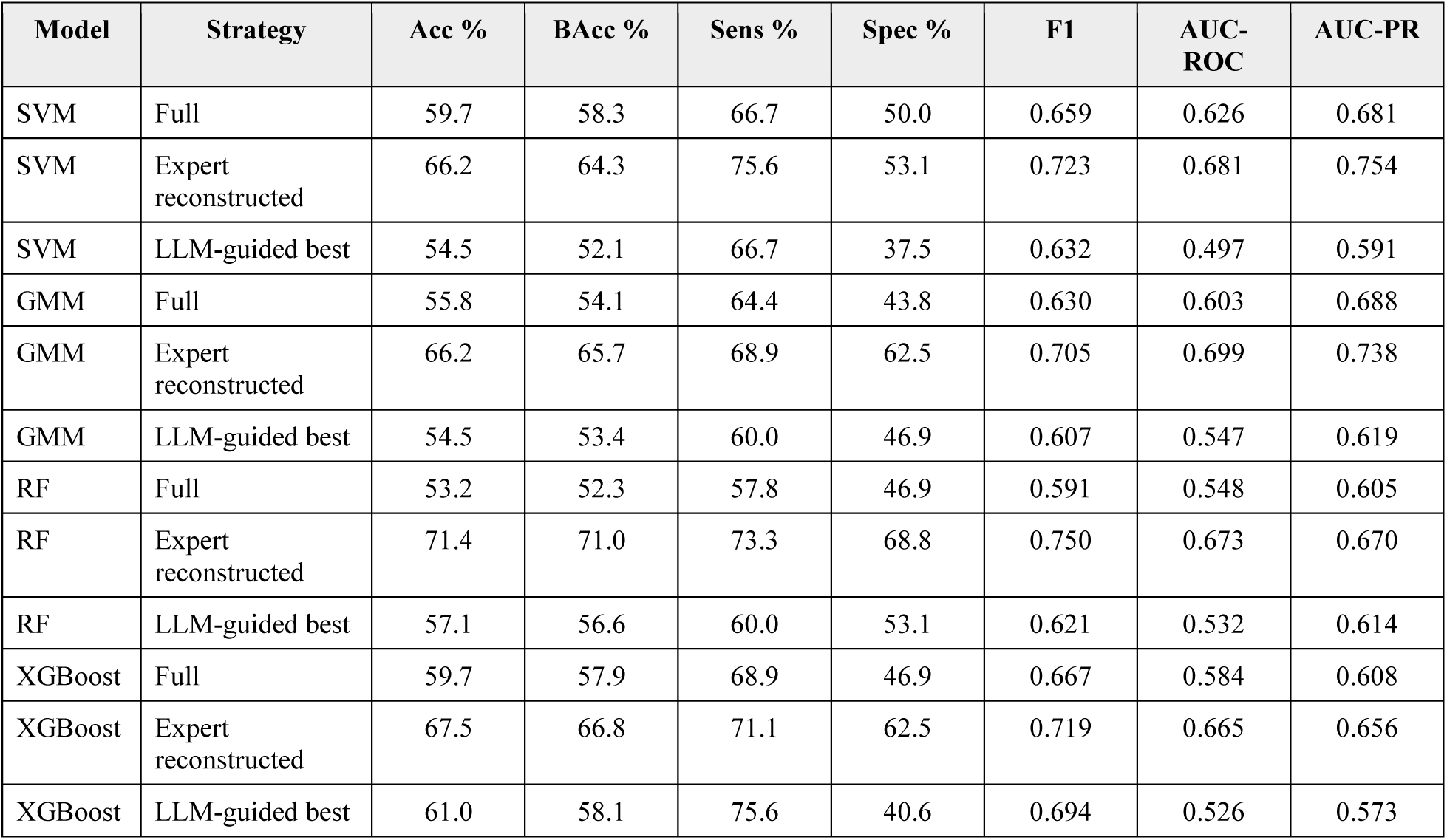
Final LOOCV performance for the controlled comparison of four classifiers × three feature strategies. All SVM rows were produced with the uncalibrated standardized-margin scoring route described in Section 2.6.1 and are reported as secondary sensitivity evidence; the primary calibrated SVM path is described in Section 3.4. For reference, the secondary elastic-net logistic regression reached 57.1% accuracy and 57.0% balanced accuracy on the expert-reconstructed strategy (Section 2.6.5). Under the pre-specified balanced-accuracy selection rule, RF/expert is the leading model; under an AUC-first rule, GMM/expert would be selected.

**Table 3b.** Primary calibrated SVM path — pre-specified pipeline outputs. Values are LOOCV performance for the pre-specified calibrated SVM pipeline (RBF SVC with class_weight=’balanced’ and train-only sigmoid calibration via CalibratedClassifierCV; inner GridSearchCV selection under F1 scoring; ANOVA k=25). The primary calibrated pipeline produced near-degenerate specificity (6–9%) and below-chance pooled AUC-ROC (0.34–0.44), reflecting hyperparameter selections that maximized ADHD-class recall at the expense of specificity under class-imbalanced inner folds. The adopted uncalibrated standardized-margin scoring route (Table 3) was selected after inspecting six scoring variants to address this collapse; see Sections 2.6.1 and 3.4.

| Strategy | Acc | Bacc | Sens | Spec | F1 | AUC-ROC | AUC-PR |
| --- | --- | --- | --- | --- | --- | --- | --- |
| Full | 50.6 | 44.7 | 80.0 | 9.4 | 0.655 | 0.336 | 0.497 |
| Expert-reconstructed | 53.2 | 46.5 | 86.7 | 6.3 | 0.684 | 0.438 | 0.548 |
| LLM-guided (best) | 54.5 | 47.6 | 88.9 | 6.3 | 0.696 | 0.335 | 0.475 |

The expert-reconstructed strategy produced the highest accuracy and balanced accuracy for every classifier. RF/expert was the overall winner by the pre-specified balanced-accuracy rule (71.0% balanced accuracy; 71.4% raw accuracy). XGBoost/expert, GMM/expert and SVM/expert followed at 67.5%, 66.2% and 66.2% accuracy, respectively. The expert strategy therefore supported a consistent domain-knowledge signal across model families rather than a single-model anomaly.

The ordering of the expert-strategy models is nevertheless metric-dependent. Although RF was selected under the pre-specified balanced-accuracy rule, it did not have the highest discrimination: GMM/expert reached AUC-ROC 0.699 and SVM/expert 0.681, against 0.673 for RF/expert. An AUC-first selection rule would therefore have selected GMM. We report this explicitly because it reinforces, rather than qualifies, the conclusion that these data do not support a stable ranking among classifiers.

The LLM-guided strategy did not outperform the expert comparator for any classifier. Relative to the full feature space it showed small gains for RF (57.1% vs 53.2%) and XGBoost (61.0% vs 59.7%), but small losses for GMM (54.5% vs 55.8%) and SVM (54.5% vs 59.7%). These results therefore show no consistent predictive advantage of LLM-guided grouping over a full-space baseline and a clear disadvantage relative to the reconstructed expert strategy.

**Figure 2.**
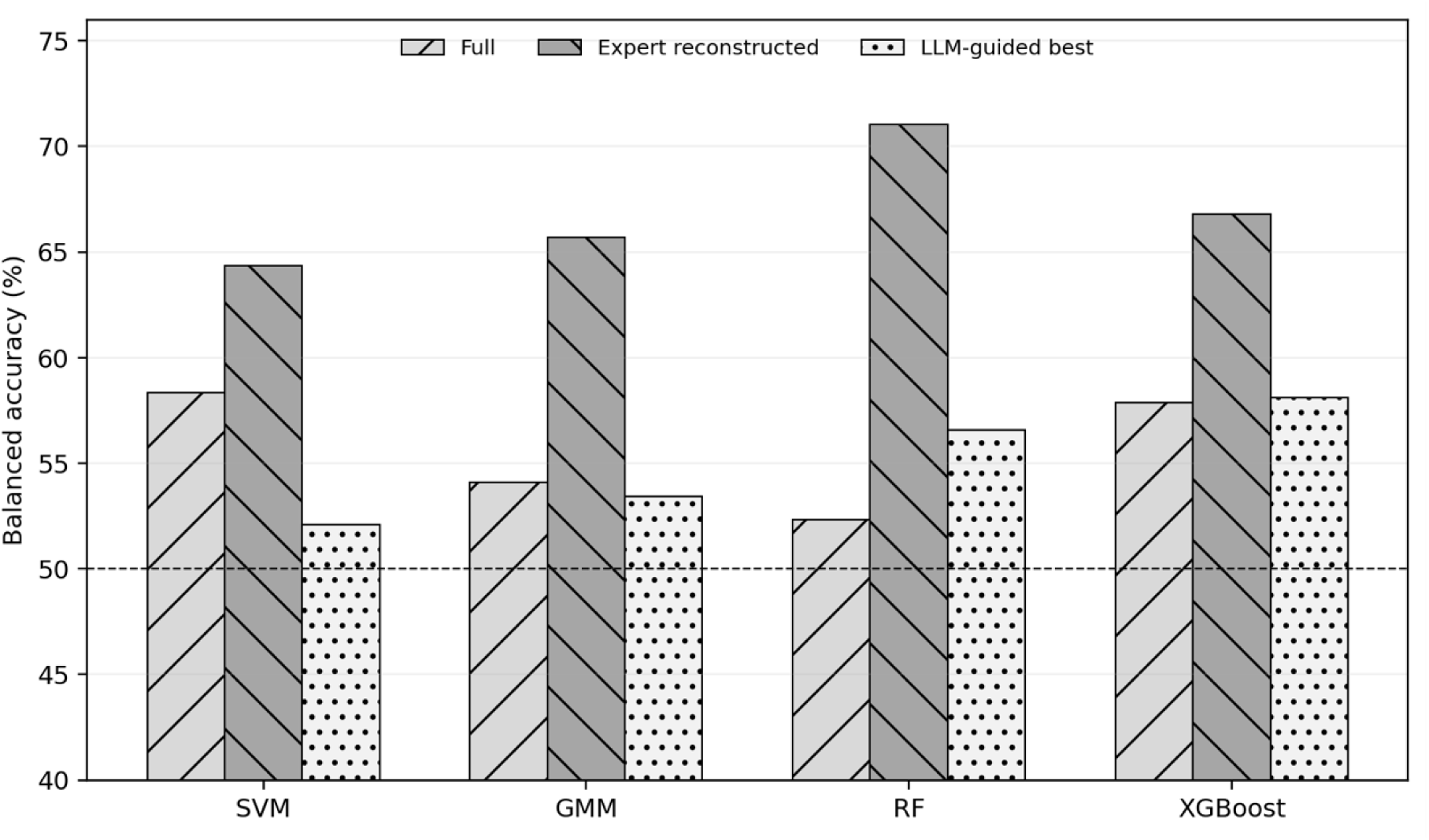
Balanced accuracy across classifiers and feature strategies under the final LOOCV assembly. The dashed line marks 50% balanced accuracy; the study no-information accuracy rate is 58.44% because classes are imbalanced.

### 3.3 Primary Random-Forest Result and Repeated-CV Robustness

RF on the 450-feature expert-reconstructed strategy correctly classified 55 of 77 children (33/45 ADHD and 22/32 controls), for accuracy 71.43% (95% Wilson CI 60.5%–80.3%) and balanced accuracy 71.04%. Sensitivity was 73.33% (95% CI 59.0%–84.0%), specificity 68.75% (95% CI 51.4%–82.0%), precision 76.74%, F1 0.750, AUC-ROC 0.673 and AUC-PR 0.670.

Across 100 repetitions of stratified 10-fold cross-validation, RF/expert mean accuracy was 62.84% (SD 3.50%; empirical 95% interval 55.84%–69.51%), mean balanced accuracy 62.32% and mean AUC-ROC 0.618. The lower repeated-CV center reinforces the uncertainty of a single LOOCV point estimate in a 77-subject cohort [17].

**Figure 3.**
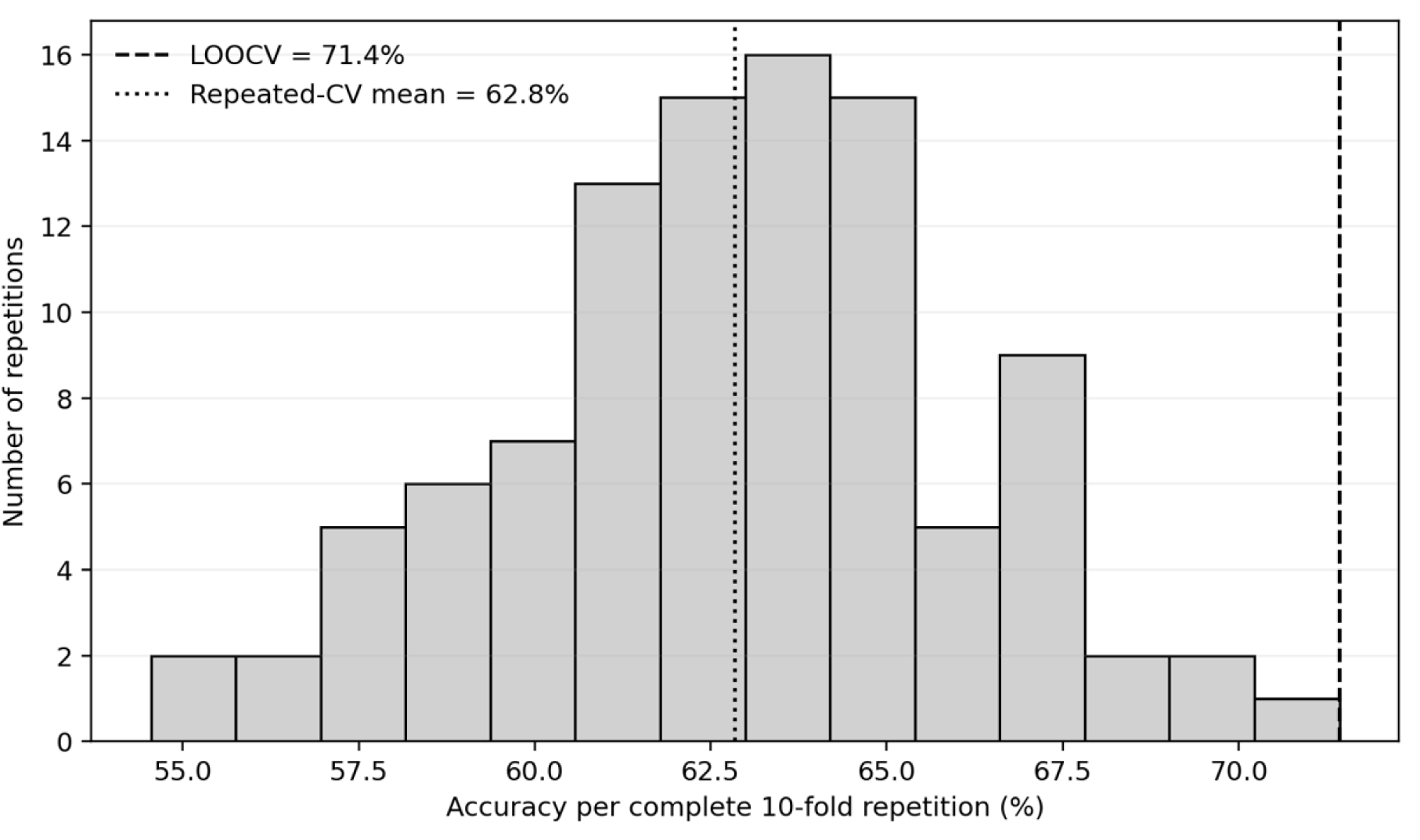
Distribution of RF/expert accuracy across 100 complete repetitions of stratified 10-fold cross-validation. Each repetition contains one out-of-fold prediction per subject.

### 3.4 Classifier Comparisons and SVM Sensitivity Path

On expert-reconstructed features, XGBoost and GMM achieved 67.53% and 66.23% accuracy, respectively. The secondary SVM sensitivity analysis also achieved 66.23% accuracy, with sensitivity 75.56%, specificity 53.13%, balanced accuracy 64.34%, F1 0.723 and AUC-ROC 0.681. The pre-specified calibrated SVM path (Table 3b/S1) produced substantially different numbers on the same folds and features: 53.2% accuracy, 86.7% sensitivity, 6.3% specificity and AUC-ROC 0.438 on the expert-reconstructed strategy, with corresponding degenerate specificity on the other two strategies (Table 3b/S1). Because the reporting route used in Table 3 was selected after a six-variant diagnostic and used a different final class-weight setting, this result is reported as secondary sensitivity evidence rather than evidence of SVM superiority; the calibrated path is included here so that the reader can assess the effect of the scoring change directly.

**Table 4.** Exact McNemar comparisons among final expert-strategy predictions.

| Model A | Model B | A-only correct | B-only correct | Raw p | Holm p |
| --- | --- | --- | --- | --- | --- |
| SVM | GMM | 9 | 9 | 1.000 | 1.000 |
| SVM | RF | 6 | 10 | 0.454 | 1.000 |
| SVM | XGBoost | 7 | 8 | 1.000 | 1.000 |
| GMM | RF | 8 | 12 | 0.503 | 1.000 |
| GMM | XGBoost | 11 | 12 | 1.000 | 1.000 |
| RF | XGBoost | 6 | 3 | 0.508 | 1.000 |

No pairwise comparison was statistically significant after Holm correction. Accordingly, the manuscript describes RF as the highest observed model rather than as statistically superior to the other classifiers.

### 3.5 Permutation, Calibration and Clinical-Utility Analyses

The available conditional fixed-configuration permutation test for RF/expert used 1,000 label permutations and yielded observed frozen-configuration LOOCV accuracy 72.73% versus a null mean of 50.70% (SD 8.44%), p = 0.006. A parallel GMM/expert test yielded observed accuracy 67.53% versus null mean 51.19% (SD 7.46%), p = 0.015. These observed accuracies differ from the nested headline cells because the permutation tests intentionally freeze a single configuration. The p-values are therefore conditional and do not correct the preceding strategy and hyperparameter search.

The permutation null means (approximately 51%) lie below the no-information rate of 58.44%. This is expected rather than anomalous: label permutation preserves the class marginals and because these pipelines use balanced class weighting or explicit class priors, the permuted-label models do not collapse to majority-class prediction. The null therefore centers near 50% rather than near the NIR and the permutation p-values should be read as tests against chance-level discrimination, not as tests against a majority-class baseline. Accuracy relative to the NIR is addressed separately by the point estimate and its Wilson interval in Section 3.3.

For RF/expert out-of-fold probabilities, the Brier score was 0.234. The calibration intercept was 0.319 (95% CI −0.146 to 0.784) and the slope 0.523 (95% CI −0.011 to 1.057), indicating correctly oriented but imperfectly calibrated probabilities. The slope below 1 is consistent with overconfident probability spread, although the confidence interval is wide.

**Figure 4.**
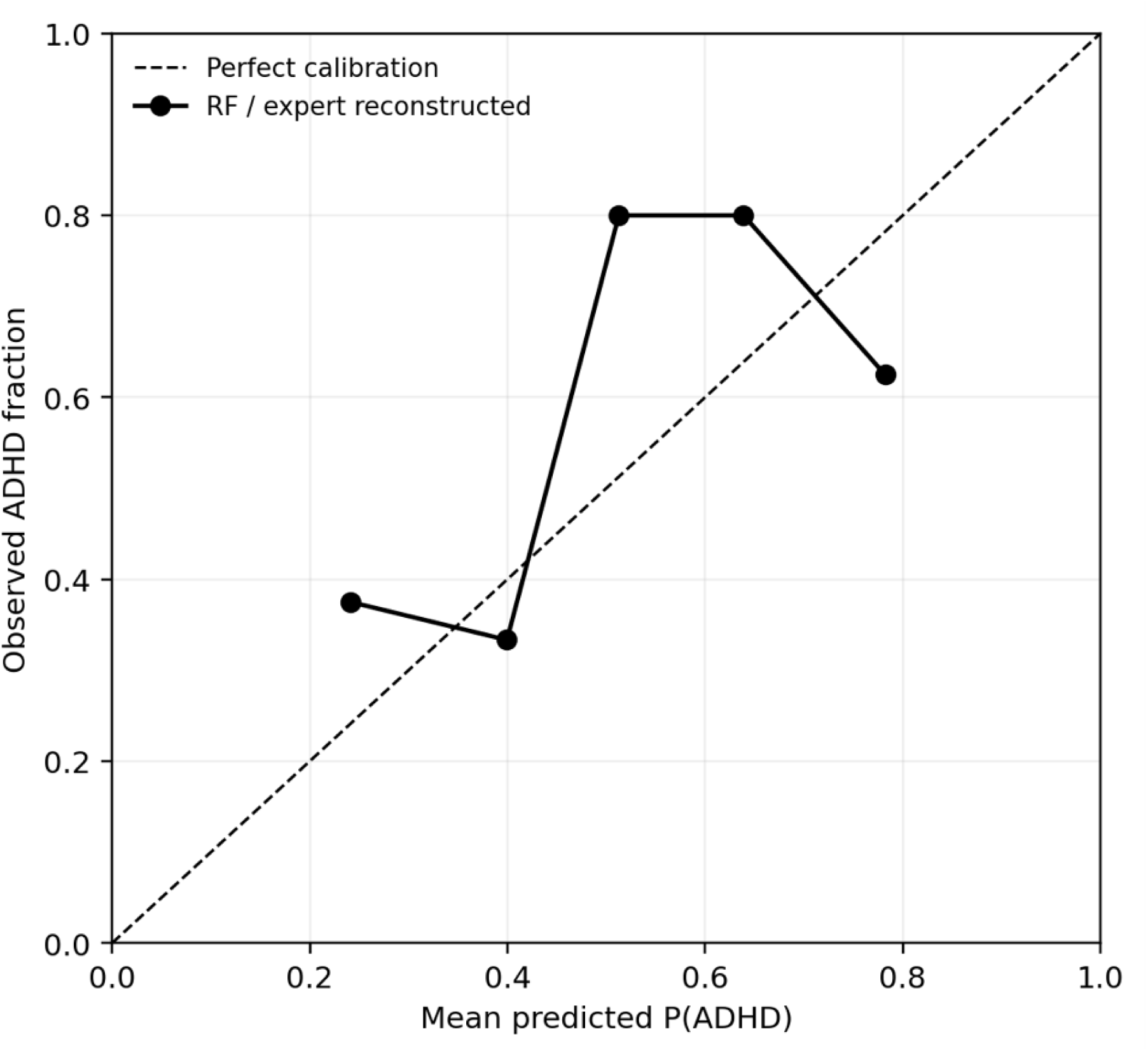
Five-bin quantile calibration curve for the primary RF/expert model using pooled out-of-fold probabilities. Calibration estimates are exploratory because of the small enriched cohort.

**Table 5.** Prevalence-aware predictive values for the primary RF/expert model.

| Assumed prevalence | PPV | NPV | LR+ | LR– | Context |
| --- | --- | --- | --- | --- | --- |
| 5.0% | 11.0% | 98.0% | 2.35 | 0.39 | Illustrative low-prevalence screening |
| 10.0% | 20.7% | 95.9% | 2.35 | 0.39 | Illustrative 10% setting |
| 30.0% | 50.1% | 85.7% | 2.35 | 0.39 | Illustrative referred setting |
| 58.4% | 76.7% | 64.7% | 2.35 | 0.39 | Study sample |

At a 5% ADHD prevalence, the observed sensitivity and specificity would imply a PPV of about 11% and an NPV of about 98%; at 10% prevalence, PPV is about 21% and NPV about 96%. These figures illustrate why the model cannot be interpreted as a stand-alone screening or diagnostic test despite accuracy above the study NIR.

### 3.6 Feature Stability and Error Overlap

Feature stability favored the expert-reconstructed strategy. A feature was counted as stable if it was selected in at least 90% of the 77 outer folds. Under this threshold and across expert-strategy folds, SVM selected 17 stable features, whereas GMM, RF and XGBoost each had 14. Eyes-closed O1 relative theta-power variability (EC_bandpower_channel_O1_theta_rel_std) was selected in 100% of folds for all four model pipelines. Other highly stable RF/expert variables included O1 and O2 theta measures, EO O1 alpha relative power, EO Pz TBR variability and VCPT O2 theta relative power. This pattern supports a reproducible posterior theta/alpha contribution, but it should not be interpreted as a validated biomarker without external replication.

Across the 12 primary model-strategy cells, five subjects were misclassified in every cell and nine were misclassified in at least 10 cells; seven of those nine were controls. The overlap suggests that some observations are consistently difficult for the present feature representation, but it does not establish an intrinsic information ceiling and is not used to justify one.

### 3.7 Sensitivity Analyses

None of the tested implementation changes satisfied the pre-stated criterion for global adoption. Log10 transformation of absolute-power features improved GMM substantially (approximately 5–11 percentage points depending on strategy) but reduced or failed to improve other models, so it was retained as a model-specific sensitivity finding rather than changing the shared primary feature matrix. XGBoost class weighting reduced performance, inner tuning of ANOVA k selected larger k values that generalized worse and replacing inner F1 with balanced accuracy did not improve the primary winner.

The 200 µV peak-to-peak artifact gate had the most important substantive effect: RF/expert accuracy fell by approximately 15.6 percentage points, from 71.4% to approximately 55.8%, which is below the 58.44% no-information rate. Artifact rejection rates also differed by group in eyes-open recordings (mean rejection approximately 0.33 for ADHD vs 0.23 for controls). One interpretation is that part of the discriminative signal available to the primary pipeline reflects movement or artifact burden or behavioral-load differences, rather than neural dynamics alone. An alternative, non-exclusive explanation is that applying the gate reduced the number of usable epochs and thereby increased noise or instability in subject-level feature estimates.

Increasing coherence resolution (nperseg 128 to 500) further reduced performance and was not adopted. These sensitivity results materially constrain the neurophysiological interpretation of the classifier.

## 4. Discussion

### 4.1 Principal Findings

Repeated stratified 10-fold cross-validation provided the more conservative estimate of RF/expert performance, with mean accuracy 62.8%, mean balanced accuracy 62.3% and mean AUC-ROC 0.618 across 100 repetitions; the empirical 95% accuracy interval was 55.8%–69.5%. The corresponding LOOCV point estimate was higher at 71.4% accuracy and 71.0% balanced accuracy and lay above the repeated-CV empirical interval. XGBoost, GMM and the secondary SVM sensitivity analysis were close behind on the expert strategy, GMM achieved the highest AUC-ROC and no pairwise classifier comparison was significant after multiplicity correction. The data therefore do not support a claim that one algorithm dominates this cohort and the identity of the leading model depends on which metric is prioritized.

The second major finding is the controlled feature-strategy result: expert-reconstructed features were strongest for every model. LLM-guided grouping never exceeded the expert comparator and showed no consistent advantage relative to the full feature space. This is a meaningful negative result because the LLM was evaluated under the same outer splits and optimization budget rather than being compared across incompatible feature spaces. The consistency of the pattern across four dissimilar model families, rather than the size of any single cell, is what supports the conclusion.

### 4.2 LLM-Guided Versus Expert Feature Engineering

The LLM component should be interpreted as structured hypothesis generation, not automated biomarker discovery. The candidate masks emphasized concepts prominent in the ADHD-EEG literature, including task-related attention, frontal spectral measures, TBR, entropy, asymmetry and coherence. Those groupings were interpretable, but literature significance is not equivalent to dataset-specific predictive value. The reconstructed expert strategy appears to have concentrated available features that were more consistently useful across models.

The negative result does not imply that LLM assistance is intrinsically unhelpful. LLMs may still reduce the burden of organizing large feature dictionaries, document candidate hypotheses or support literature mapping. The present result is narrower: with these features, this cohort, these seven candidate masks and leakage-controlled subject-level evaluation, LLM-guided grouping did not improve predictive performance over expert reconstruction. Future LLM studies should prospectively freeze the model and version, the prompt, the candidate masks and the evaluation plan before any performance is examined.

### 4.3 Classifier Comparison, Selection Bias and Uncertainty

Post-hoc feature and configuration selection can produce optimistic performance estimates in small datasets. For this reason, the primary conclusions are based on the frozen fold-contained evaluation framework, while analyses involving post-hoc scoring diagnostics are reported only as secondary sensitivity results.

The SVM scoring diagnostic also illustrates why metric construction matters. Train-only sigmoid calibration produced extremely low specificity and inverted the pooled ranking, whereas uncalibrated margins recovered a more plausible discrimination signal. Because the adopted uncalibrated standardized-margin route was selected after inspecting six scoring variants and refits the model with a different class-weight setting, it is reported transparently as a secondary sensitivity path rather than allowed to drive the headline narrative.

The secondary elastic-net logistic-regression reference achieved 57.1% accuracy and 57.0% balanced accuracy on the expert-reconstructed strategy. Its accuracy was slightly below the 58.44% no-information rate, providing no evidence of useful linear discrimination under this reference specification.

More generally, the 77-subject sample is too small to interpret modest differences among classifiers as a stable ordering. Varoquaux [17] showed that cross-validation uncertainty can be large at neuroimaging sample sizes of this magnitude. The repeated-CV shift from 71.4% LOOCV to 62.8% mean accuracy for the primary model is a practical demonstration of that uncertainty.

### 4.4 Comparison With Prior EEG-ADHD Studies

Rohani et al. [22] reported 75.8% accuracy for a linear SVM using an expert-selected 113-feature EEG/ERP/behavioral representation on the 103-subject source dataset. Direct numerical comparison is not appropriate because the present 450-feature expert reconstruction lacks reaction-time, omission and commission and true ERP peak features, uses a smaller frozen cohort and employs a different model-selection framework.

Kim et al. [30] provide an important contemporary subject-level benchmark. In 168 participants (107 ADHD, 61 neurotypical), they used 19-channel EEG, 30-second segmentation, XGBoost and leave-one-subject-out validation, reporting 90.81% accuracy and F1 = 0.9347. Their segmentation should not be equated with leakage because the evaluation unit remained subject-separated. The large performance gap relative to our 71.4% may reflect the larger cohort, different signal processing and feature representation, segmentation and augmentation design, cohort composition and other study-specific factors. Cross-study accuracy differences cannot identify a single cause.

García-Ponsoda et al. [31] further demonstrated that EEG preprocessing and temporal segmentation can materially affect ADHD-classification performance, reinforcing the importance of evaluating preprocessing choices and maintaining appropriate subject-level separation when segmented EEG data are used.

The more precise conclusion is that the evaluation level must be examined study by study. Segment-level random splits can create subject overlap, whereas segmentation combined with genuine subject-level separation, as in Kim et al. [30], can remain methodologically valid. Supplementary Table S7 makes this distinction explicit.

### 4.5 Clinical Interpretation

This study addresses ADHD-versus-healthy-control discrimination, not clinical differential diagnosis. Real diagnostic settings include anxiety, autism spectrum conditions, learning disorders, sleep problems, mood disorders, medication effects and other sources of EEG variation. The source cohort is valuable because diagnoses were clinically established, IQ screening was used and children were medication-free at recording, but the binary control comparison remains simpler than clinical practice.

The RF/expert likelihood ratios (LR+ 2.35; LR− 0.39) represent only modest shifts in probability. The prevalence analysis is especially instructive: at 5–10% prevalence, PPV is approximately 11–21% even with NPV around 96–98%. Such a profile may be more compatible with exploratory risk stratification than definitive diagnosis and even that interpretation requires external calibration in a representative population. We therefore make no claim of clinical readiness.

### 4.6 Limitations

Several limitations are central to interpretation. First, n = 77 is small for multivariable machine learning and yields wide uncertainty and there is no independent external test cohort. Second, attrition was strongly differential (49 to 45 ADHD versus 54 to 32 controls), yet participant-level reasons and demographic characteristics for included versus excluded subjects were not recoverable. A later file audit also identified six additional cleaned ADHD recordings outside the frozen matrix; retaining the frozen cohort was methodologically preferable to post-hoc expansion, but the discrepancy requires prospective resolution in future work.

Third, the expert-reconstructed strategy is not the original Rohani 113-feature vector. Behavioral reaction time, omission and commission errors, arousal-index variables and true ERP amplitude and latency features were unavailable in the primary feature universe, limiting both replication and biological comparability. Fourth, the artifact-gating sensitivity analysis substantially reduced RF performance and revealed group differences in artifact rejection rates. Some predictive signal may therefore reflect movement or task-engagement burden rather than purely neural physiology. At the same time, the reduction in performance may partly reflect loss of usable epochs and less stable feature estimation after stricter artifact rejection. The artifact-gating result should therefore be interpreted as evidence of sensitivity to preprocessing choices, rather than as proof that the original classifier was driven by artifact burden.

Fifth, the final SVM uncalibrated standardized-margin route followed a six-variant diagnostic and changes the class-weight setting relative to the frozen inner search; it should be viewed as secondary. Sixth, the available permutation p-values are conditional on frozen selected configurations and do not constitute full-procedure, search-adjusted permutation tests. Seventh, calibration and decision-curve analyses are based on only 77 out-of-fold observations from an enriched case-control-like sample and should not be transported directly to population screening. Eighth, the leading model was selected by a balanced-accuracy rule and would not have been selected under an AUC-first rule. Finally, this is a healthy-control comparison and does not establish specificity against clinically relevant differential diagnoses.

### 4.7 Future Work

The next study should freeze cohort inclusion, feature masks and the full analysis plan before outcome inspection; retain complete participant metadata and preprocessing provenance; and evaluate the final pipeline in an independent pediatric cohort. Re-access to raw ERP event markers would permit reconstruction of the true ERP amplitude and latency and behavioral variables that were unavailable here.

Artifact handling should be standardized prospectively, with movement burden quantified as a separate covariate rather than inadvertently embedded in the classifier. A direct test of this confound, modelling diagnostic group from the per-subject epoch-rejection rate alone under the same subject-level splits, would establish how much of the observed discrimination is attributable to artifact burden rather than to spectral or complexity features and should be reported alongside the primary model in any replication. A direct extension of the present work is to repeat the full pipeline on the 83-subject audited cohort, including the six additional cleaned ADHD recordings and to report it as a pre-declared cohort-definition sensitivity analysis; demonstrating stability across the two cohort definitions would be stronger evidence than retaining the frozen matrix alone. Larger multicenter samples should include clinically realistic comparison groups and support external calibration, decision-curve analysis and subgroup assessment.

## 5. Conclusion

Across 100 repetitions of stratified 10-fold cross-validation, RF using the expert-reconstructed EEG strategy achieved mean accuracy 62.8%, mean balanced accuracy 62.3% and mean AUC-ROC 0.618 (empirical 95% accuracy interval 55.8%–69.5%). The corresponding LOOCV point estimate was higher at 71.4% accuracy and 71.0% balanced accuracy. RF was selected under the pre-specified balanced-accuracy rule, whereas GMM/expert achieved the highest AUC-ROC (0.699) and pairwise classifier differences were not statistically significant after Holm correction. A 200 µV artifact gate reduced RF/expert accuracy to approximately 55.8%, below the 58.4% no-information rate; this sensitivity may reflect both removal of artifact-related signal and loss of usable epochs. LLM-guided feature grouping never outperformed the reconstructed expert strategy and showed no consistent advantage over the full feature space. These findings support transparent benchmarking under strict validation rather than claims of LLM or algorithmic superiority. External validation and improved cohort and preprocessing provenance are required before clinical interpretation.

## Data Availability

Analysis code and frozen reproducibility materials (per-subject out-of-fold predictions, fold-specific selected-feature and hyperparameter logs, software-version records and verification scripts) are publicly available on the Open Science Framework: https://osf.io/fs89b/ . The EEG dataset was originally collected by Rohani et al. (2022, doi:10.22067/cke.2022.70922.1014); access to raw data is subject to the terms of the original data holders.

## Acknowledgments

The authors thank Andreas Mueller (Brain and Trauma Foundation, Chur, Switzerland; HBImed AG, Chur, Switzerland) for detailed pre-submission review of the manuscript and methodology and the Motor Behavior Laboratory at Ferdowsi University of Mashhad, Iran, for providing access to the EEG dataset used in this study. The authors also acknowledge the use of a large language model as a feature-engineering assistant, as described in Section 2.4.3 and Supplementary Methods S2. The model was not used to generate, analyze or interpret participant data and it had no access to diagnostic labels, raw EEG or performance results.

## Author Contributions

All authors contributed to the conceptualization, methodology, investigation, data curation, formal analysis, validation, interpretation of results and review and editing of the manuscript. Faezeh Rohani, first author of the source study [22], is a co-author of the present secondary analysis; she provided the original dataset and associated resources and contributed to the final verification of the study materials and results. All authors have reviewed and approved the final manuscript.

## Ethics Statement

This secondary analysis used previously collected and fully anonymized data. The source study was approved by the Research Ethics Committee of Islamic Azad University, Lahijan Branch, Iran (approval no. IR.IAU.LIAU.REC.1398.013, approved 6 August 2019), and the parents of all participants provided written informed consent.

The present work is a secondary analysis of fully anonymized data involving no participant contact and no new data collection. No identifiable participant data were accessible to the present authors at any stage.

## Data and Code Availability

Analysis code and frozen reproducibility materials are publicly available on the Open Science Framework at https://osf.io/fs89b/ (see also Supplementary Results S6). The EEG dataset was originally collected by Rohani et al. [22]; access to raw data is subject to the terms of the original data holders. The analysis is reproducible from the frozen 77-subject feature matrix, per-subject out-of-fold predictions, fold-specific selected-feature and hyperparameter logs, software-version records and verification scripts.

## Reporting Guideline

The manuscript was prepared with reference to the TRIPOD+AI reporting recommendations for prediction-model studies [29]. A reporting crosswalk is provided in the Supplementary Material, and a completed TRIPOD+AI checklist is available from the authors.

## Supplementary Material

### Supplementary Methods S1. Registered Feature Strategies

| Strategy/mask | Candidate N | Definition |
| --- | --- | --- |
| Full | 1,185 | All valid gamma-free features |
| Expert reconstructed | 450 | TBR, EO/EC coherence, available theta/alpha/beta channel band power |
| LLM: VCPT-only | 397 | VCPT-condition features |
| LLM: frontal-all | 324 | Frontal feature families |
| LLM: VCPT + frontal + TBR | 424 | Task + frontal + TBR |
| LLM: global + asymmetry + coherence | 414 | Global spectral, asymmetry, coherence |
| LLM: entropy + Hjorth + time | 222 | Complexity and time-domain |
| LLM: EO + EC only | 788 | Resting-state features |
| LLM: VCPT + entropy + TBR | 505 | Task + complexity + TBR |

For llm_best, the mask was selected inside outer-training data by inner 5-fold cross-validation. The LLM had no access to participant labels, raw EEG, split membership or performance results.

### Supplementary Methods S2. Reconstructed LLM Prompt Specification

#### Original LLM provenance

The retained project records confirm that an LLM was used as a literature-informed feature-engineering assistant, but they do not preserve a verifiable record of the provider, exact model/version, date of interaction or verbatim original prompt. Accordingly, these details cannot be reported retrospectively with certainty.

#### Information available to the LLM

EEG feature names and metadata, recording conditions (EO, EC, VCPT), channel information, feature families and established ADHD-EEG concepts.

#### Information withheld from the LLM

participant identifiers, ADHD/control labels, raw EEG recordings, train/test assignments, cross-validation outcomes, classifier performance and final model results.

#### Reconstructed task specification

“You are assisting with feature engineering for a pediatric ADHD EEG classification study. You are not given participant labels, raw EEG recordings, train/test splits or model-performance results. Based only on the available EEG feature metadata, recording conditions, channel labels and established ADHD-EEG literature organize the available features into neurophysiologically meaningful candidate subsets for subsequent machine-learning evaluation. Consider feature families related to attentional control, response inhibition, frontal spectral activity, theta/beta ratio, spectral power, entropy/complexity, hemispheric asymmetry, coherence and task-related EEG measures. The output should define candidate feature groups rather than select individual participants or make diagnostic predictions.”

This reconstructed task specification is based on the preserved methodological records and should not be interpreted as the verbatim original prompt used during the initial LLM interaction.

### Supplementary Results S1. Full Repeated 10-Fold x100 Robustness Table

| Model | Strategy | Accuracy mean +/- SD % | Empirical 95% interval % | Mean BAcc % | Mean AUC |
| --- | --- | --- | --- | --- | --- |
| SVM | Full | 56.77 +/- 2.73 | 51.95-61.72 | 49.46 | 0.476 |
| SVM | Expert reconstructed | 56.81 +/- 2.67 | 50.65-61.04 | 49.76 | 0.505 |
| SVM | LLM-guided best | 56.44 +/- 3.11 | 50.65-61.72 | 49.35 | 0.471 |
| GMM | Full | 58.97 +/- 4.77 | 50.65-68.21 | 57.79 | 0.602 |
| GMM | Expert reconstructed | 60.97 +/- 4.96 | 51.95-70.81 | 60.35 | 0.629 |
| GMM | LLM-guided best | 55.90 +/- 4.90 | 46.75-64.32 | 54.06 | 0.559 |
| RF | Full | 54.68 +/- 4.30 | 48.05-62.34 | 54.11 | 0.536 |
| RF | Expert reconstructed | 62.84 +/- 3.50 | 55.84-69.51 | 62.32 | 0.618 |
| RF | LLM-guided best | 53.25 +/- 4.31 | 43.47-59.74 | 52.20 | 0.512 |
| XGBoost | Full | 55.94 +/- 4.02 | 47.37-63.02 | 54.63 | 0.532 |
| XGBoost | Expert reconstructed | 62.26 +/- 3.91 | 55.84-69.51 | 60.93 | 0.607 |
| XGBoost | LLM-guided best | 56.31 +/- 4.61 | 48.05-63.64 | 54.19 | 0.523 |

Important SVM note: the frozen repeated-CV batch predates adoption of the v4 dec_none_z SVM reporting path and therefore reflects the original calibrated SVM construction. These SVM repeated-CV values are presented for provenance only and are not used to validate the final 66.2% dec_none_z LOOCV result.

### Supplementary Results S2. Conditional Permutation Tests

| Model | Strategy | Permutations | Observed fixed config | Null mean | Null SD | p (+1) |
| --- | --- | --- | --- | --- | --- | --- |
| RF | Expert reconstructed | 1,000 | 72.73% | 50.70% | 8.44% | 0.006 |
| GMM | Expert reconstructed | 1,000 | 67.53% | 51.19% | 7.46% | 0.015 |

These tests freeze a configuration selected from real-label analyses. They therefore test whether that fixed configuration performs above a permuted-label null, but do not correct for the preceding strategy/hyperparameter search. The nested LOOCV cells in the main manuscript remain the headline performance estimates.

### Supplementary Results S3. Sensitivity-Analysis Summary

| Sensitivity change | Decision | Observed implication |
| --- | --- | --- |
| Log10 absolute-power transform | Not adopted globally | GMM improved by approximately 5-11 pp across strategies; other models neutral/worse. |
| XGBoost scale_pos_weight=32/45 | Not adopted | Reduced XGBoost balanced accuracy, including expert strategy. |
| Tune ANOVA k from 10-40 | Not adopted | All four expert models tied or worsened; larger k was frequently selected. |
| Inner metric F1 -> balanced accuracy | Not adopted | No improvement in primary RF; other models mixed/slightly worse. |
| 200 microvolt peak-to-peak epoch gate | Not adopted; key limitation | RF/expert accuracy decreased about 15.6 pp; EO rejection rate was higher in ADHD (about .36) than controls (about .23). |
| Coherence nperseg 128 -> 500 | Not adopted | Reduced performance across most cells. |

### Supplementary Results S4. Feature Stability and Error Overlap

| Model | Stable (predefined threshold) | Moderate |
| --- | --- | --- |
| SVM | 17 | 7 |
| GMM | 14 | 5 |
| RF | 14 | 5 |
| XGB | 14 | 5 |

The feature EC_bandpower_channel_O1_theta_rel_std was selected in 100% of expert-strategy folds for all four model pipelines. Across all 12 primary cells, five subjects were misclassified in every cell; nine were misclassified in at least 10 cells, including seven controls and two ADHD participants.

### Supplementary Results S5. Calibration and Decision Analysis

RF/expert Brier score = 0.234; calibration intercept = 0.319 (95% CI −0.146 to 0.784); calibration slope = 0.523 (95% CI −0.011 to 1.057). Decision-curve analysis is exploratory because the study sample is enriched for ADHD and does not represent a target-population prevalence.

**Supplementary Figure S1.**
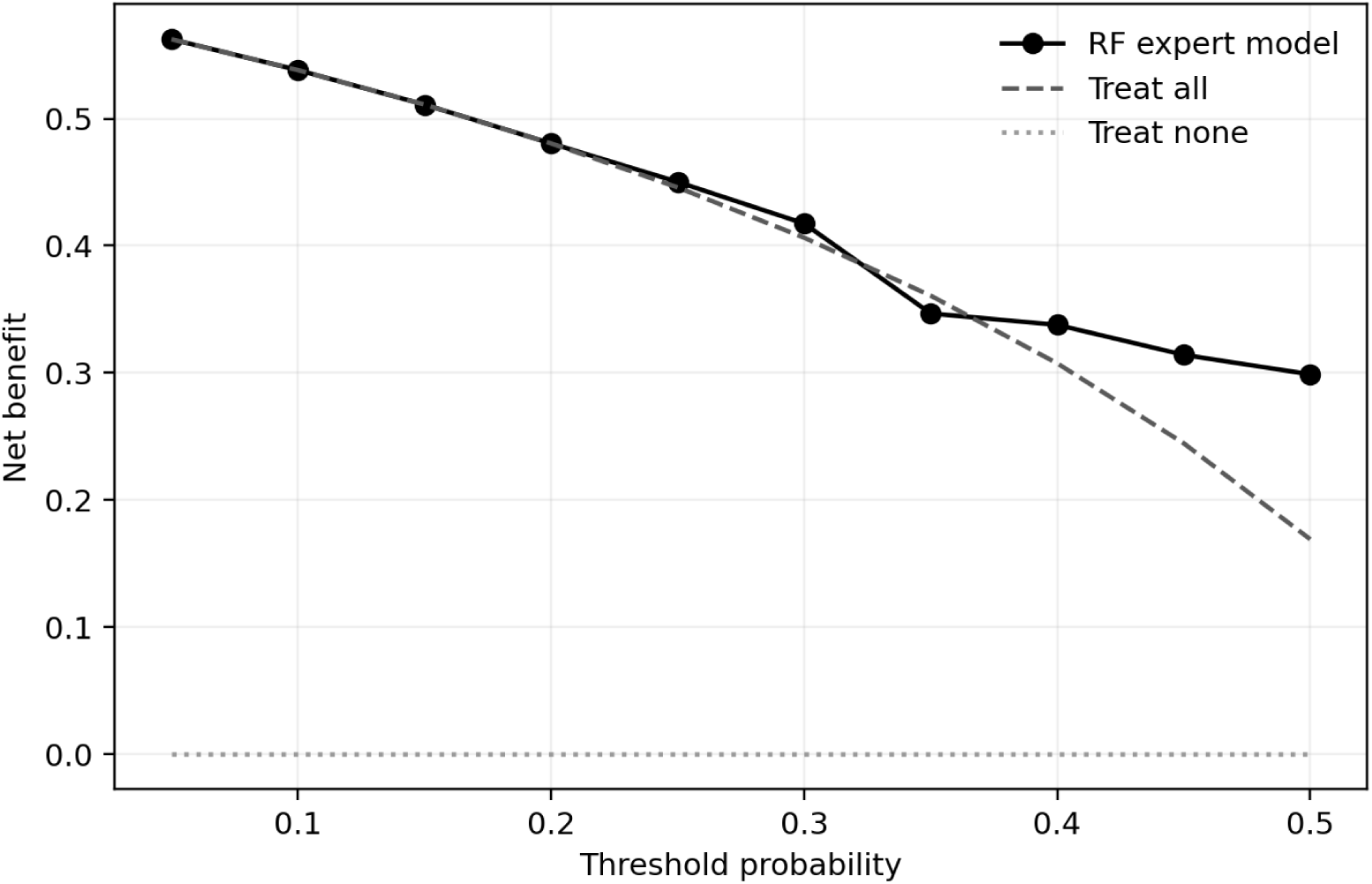
Exploratory sample-based decision curve for RF/expert.

### Supplementary Results S6. Software and Reproducibility

| Component | Version / environment |
| --- | --- |
| numpy | 2.5.2 |
| pandas | 3.0.5 |
| platform | Windows-11-10.0.26200-SP0 |
| python | 3.12.5 |
| mne | 1.12.1 |
| scikit_learn | 1.9.0 |
| scipy | 1.18.0 |
| xgboost | 3.4.1 |

Random seed base: 42. Training loops used n_jobs=1. The result package stores per-subject predictions, selected features by fold, hyperparameters by fold, software versions and integrity-verification outputs. The v4 verifier recomputes reported metrics from raw predictions.

#### Code and reproducibility materials

The analysis code and frozen reproducibility materials used in this study are available through the Open Science Framework (OSF) at: https://osf.io/fs89b/. The repository contains the analysis scripts, configuration files, per-subject predictions, fold-specific selected-feature and hyperparameter records, software-version information and verification outputs supporting the analyses reported in this manuscript.

**Supplementary Table S7.** Evaluation-Scheme Context for Key Comparator Studies.

| Study | N | Representation | Evaluation unit | Reported result | Interpretive note |
| --- | --- | --- | --- | --- | --- |
| Rohani et al. 2022 [1] | 103 | EEG/ERP + behavior | Cross-validation in source study; different feature set | 75.8% linear SVM reported | Original source cohort; direct numeric comparison limited. |
| Kim et al. 2025 [4] | 168 (107 ADHD/61 NT) | 19-channel EEG; 30-s segments | Leave-one-subject-out | 90.81% XGBoost | Segmentation plus LOSO is not automatically identity leakage because subjects remain separated. |
| Present study | 77 (45 ADHD/32 control) | Handcrafted EEG subject features | Nested outer LOOCV; repeated 10-fold x100 | 71.4% RF/expert LOOCV; 62.8% repeated-CV mean | All supervised selection inside outer-training data; no external validation. |
| García-Ponsoda et al. 2024 [31] | 121 (61 ADHD/60 TD) | 19-channel EEG; preprocessing variants and temporal segmentation | Subject-level 5-fold cross-validation | Up to 86.1% accuracy; performance decreased with stronger artifact removal | Shows that preprocessing and temporal segmentation materially affect reported accuracy; subject-level evaluation avoids identity leakage despite segmentation. |

### Supplementary Reporting Crosswalk: TRIPOD+AI-Oriented Items

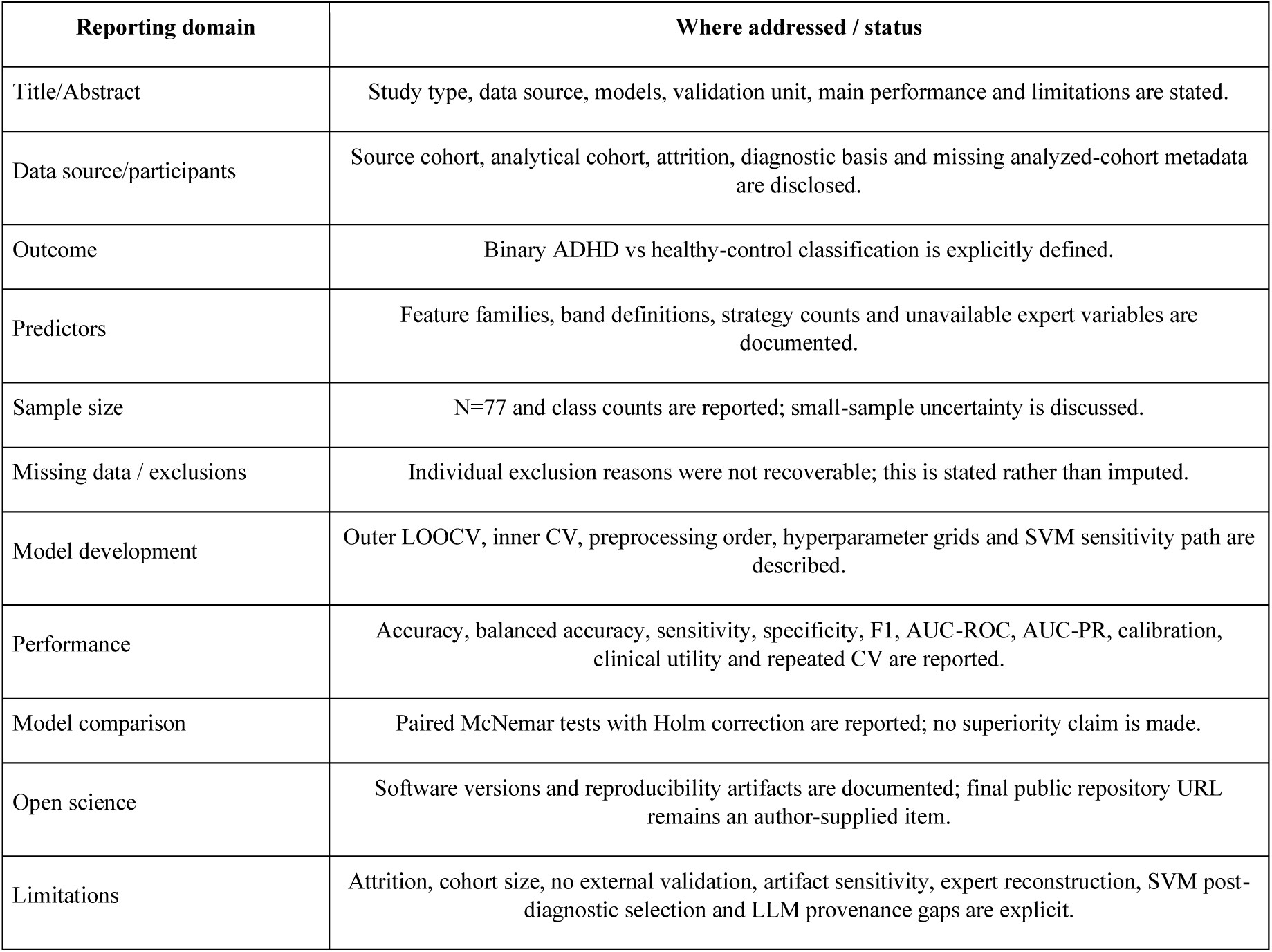

The 1,185-feature primary matrix comprises 197 unique feature definitions per epoch, each computed under three recording conditions (EO, EC, VCPT) and aggregated across epochs by mean and standard deviation (197 × 3 conditions × 2 aggregations = 1,182, plus 3 VCPT-specific stimulus counts = 1,185). Feature families use different channel subsets and different spatial scopes as described below.

Full 19-channel montage available in cleaned EDFs: Fp1, Fp2, F3, F4, F7, F8, Fz, C3, C4, Cz, T3, T4, T5, T6, P3, P4, Pz, O1, O2.

Reduced 13-channel subset used by TBR, entropy, and per-channel band-power families: Fp1, Fp2, F3, F4, Fz, C3, C4, Cz, P3, P4, Pz, O1, O2 (excludes lateral frontal F7/F8 and temporal chain T3/T4/T5/T6). This subset is applied consistently across all conditions.

Regional groupings used by band-power and asymmetry families:

frontal: Fp1, Fp2, F3, F4, F7, F8, Fz

central: C3, C4, Cz

parietal: P3, P4, Pz

occipital: O1, O2

temporal: T3, T4, T5, T6

**Supplementary Table S8.** Feature Structure.

| Family | Per-epoch count | Scope | Channels / pairs used | Bands / measures | Per-condition count | Total in matrix |
| --- | --- | --- | --- | --- | --- | --- |
| **Band power** | **129** | mixed | see sub-rows | delta, theta, alpha, beta | 129 | 774 |
| — channel-level | 78 | 13 channels | Fp1, Fp2, F3, F4, Fz, C3, C4, Cz, P3, P4, Pz, O1, O2 | theta, alpha, beta (abs + rel each = 6 per channel); delta not computed per channel | 78 | 468 |
| — region-level | 40 | 5 regions | frontal, central, parietal, occipital, temporal | delta, theta, alpha, beta (abs + rel each = 8 per region) | 40 | 240 |
| — global (whole-scalp) | 11 | 1 global | scalp-averaged | delta abs/rel (2); theta abs/rel/alpha-ratio/beta-ratio (4); alpha abs/rel/beta-ratio (3); beta abs/rel (2) | 11 | 66 |
| **Entropy** | **27** | 13 channels + 1 global | Fp1, Fp2, F3, F4, Fz, C3, C4, Cz, P3, P4, Pz, O1, O2 (approximate + sample per channel = 26); global Shannon entropy of amplitude histogram (1) | approximate, sample, shannon | 27 | 162 |
| **Theta-beta ratio (TBR)** | **13** | 13 channels | Fp1, Fp2, F3, F4, Fz, C3, C4, Cz, P3, P4, Pz, O1, O2 | one TBR value per channel | 13 | 78 |
| **Coherence** | **12** | 4 homologous pairs | Fp1-Fp2 (implicit via asymmetry family — see below), F3-F4, C3-C4, P3-P4, O1-O2 (4 pairs) | coherence in theta, alpha, beta bands (3 bands × 4 pairs) | 12 | 72 |
| **Time-domain** | **7** | 1 global | scalp-averaged | mean, standard deviation, RMS, skewness, kurtosis, Shannon entropy of amplitude histogram, zero-crossing rate | 7 | 42 |
| **Asymmetry** | **6** | 2 homologous pairs | Fp1-Fp2, F3-F4 | ln(right/left) power in theta, alpha, beta bands (3 bands × 2 pairs) | 6 | 36 |
| **Hjorth parameters** | **3** | 1 global | scalp-averaged | activity, mobility, complexity | 3 | 18 |
| **VCPT stimulus counts** | **3 (VCPT only)** | task-conditional | — | Go/NoGo/false-alarm counts | 3 (VCPT only) | 3 |
| **Total** | **197 per epoch** |  |  |  | **197 × 3 conditions = 591** | **1,185** |

Aggregation across epochs: for each per-epoch feature and each condition, mean and standard deviation across the subject’s available epochs are added to the subject-level matrix, producing 2 × 197 × 3 = 1,182 features per subject, plus 3 VCPT-only stimulus counts (single value per subject), for the 1,185-feature primary matrix.

The mismatch between the 19-channel available montage and the 13-channel subset used by TBR, entropy, and per-channel band-power was inherited from the feature-extraction pipeline (fe_svm_subject.py). This channel subset is applied uniformly across conditions and classifiers; the excluded six channels (F7, F8, T3, T4, T5, T6) do contribute to region-and global-level features (frontal region uses F7/F8; temporal region uses T3/T4/T5/T6).

